# Lymphatic filariasis antigen and microfilaria epidemiology in Samoa in 2019, six months post triple-drug mass drug administration

**DOI:** 10.1101/2024.08.26.24312592

**Authors:** Helen J Mayfield, Harriet Lawford, Benn Sartorius, Patricia M Graves, Sarah Sheridan, Terese Kearns, Shannon M. Hedtke, Katherine Gass, Take Naseri, Robert Thomsen, Colleen L Lau

## Abstract

**Background:** Elimination of lymphatic filariasis (LF) as a public health problem remains an ongoing challenge in the Pacific region. In 2018, Samoa was the first country to implement a national triple-drug mass drug administration (MDA) using ivermectin, diethylcarbamazine, and albendazole (IDA). This study reports on antigen (Ag) and microfilaria (Mf) prevalence in Samoa in 2019, 6-8 months post-MDA, and evaluates the effectiveness of the intervention in reducing Ag prevalence to below a 2% threshold. We also compared the change in Ag prevalence pre- and post-MDA between 5-9-year-olds and ≥10-year-olds to assess the suitability of young children as the target group for transmission assessment surveys (TAS).

**Methodology:** We surveyed 30 randomly selected and 5 purposefully selected primary sampling units (PSUs) in Samoa in 2018 (1.5-3.5 months post triple-drug MDA) and 2019 (6-8 months post triple-drug MDA). In each PSU, we conducted a community survey of 15-20 randomly selected households and a convenience survey of 5-9-year-old children. Demographic details were collected using an electronic questionnaire. A finger prick blood sample was collected from all participants to test for Ag and Mf. Prevalence estimates were adjusted for age, sex, and survey design.

**Principal Findings:** There was no significant change in adjusted Ag prevalence in the 30 randomly selected PSUs between 2018 (3.9% [95% CI: 2.7-5.6%]) and 2019 (4.1% [95% CI 2.7-5.9%]). In these PSUs, significantly higher Ag prevalence was observed in participants aged ≥10 years (4.6%, 95% CIs 3.0-6.7%) compared to 5-9-year-olds (1.1%, 95% CIs 0.5-2.2%).

**Conclusions/Significance:** A single round of triple-drug MDA was insufficient to break LF transmission in Samoa 6-8 months post-MDA. Significantly higher Ag prevalence in participants ≥10 years old also supports existing evidence that basing elimination thresholds on Ag prevalence among 6-7-year-olds may not be the most suitable strategy for post-MDA surveillance.

**Author Summary:** Elimination of lymphatic filariasis (LF) as a public health problem remains an ongoing challenge in many countries in the Pacific. Mass drug administration (MDA) is used to treat at-risk populations at the regional or national level with the aim of breaking transmission between humans and mosquitoes. In 2018, Samoa was the first country to distribute a national triple-drug MDA, combining ivermectin, diethylcarbamazine, and albendazole. In this study we compared the prevalence and clustering of antigen (Ag) and microfilaria (Mf) pre- and post-MDA. We observed evidence of ongoing LF transmission in Samoa 6-8 months post triple-drug MDA, with an estimated antigen prevalence in 30 randomly selected primary sampling units of 4.1% (95% CI 2.7-5.9%), and Mf-positive participants identified in all four administrative regions. This supports current guidance from the World Health Organization that multiple intervention rounds are needed to break the transmission cycle between humans and mosquitoes. Higher Ag prevalence in those aged ≥10 years, suggests that surveillance of adults rather than children should be used to inform programmatic decisions for disease elimination. Based on the significant Ag clustering at the household level, targeted surveillance and treatment of household members of Ag-positive people is recommended.

## Introduction

Lymphatic filariasis (LF) is a vector-borne disease that can cause severe physical disfigurement in the form of irreversible lymphedema and scrotal hydrocoeles, contributing to mental health issues, social stigmatisation and economic inequity [1]. The Global Program to Eliminate LF (GPELF) aims to eliminate LF as a public health problem by interrupting transmission to prevent new infections and managing morbidity to improve the well-being and outcomes for those living with LF-related lymphedema. There have been notable successes with 17 endemic countries achieving elimination as a public health problem [2] and delivering over 8.6 billion treatments through GPELF between 2000 and 2020 [3].

The primary intervention strategy for LF elimination involves wide-scale treatment of the at-risk population through mass drug administration (MDA) programmes [4]. The World Health Organization (WHO) Neglected Tropical Diseases Roadmap 2030 [2] sets global milestones and targets to prevent, control, eliminate, and eradicate 20 neglected tropical diseases (NTDs), with this number growing to 21 in December 2023 [5]. To achieve the goal of eliminating LF as a public health problem, WHO recommends all LF endemic districts implement multiple rounds of high-coverage MDA, followed by post-treatment and, ultimately, post-validation surveillance. In addition, WHO requires all endemic areas to implement a minimum package of care for LF morbidity by 2030 [6].

Most commonly, MDA programs for LF elimination include multiple rounds of albendazole with either diethylcarbamazine (DEC) or ivermectin [1]. For countries where elimination has not been achieved with two-drug MDA, the WHO now recommends that ivermectin be distributed alongside DEC and albendazole (i.e. triple-drug MDA) for areas without endemic onchocerciasis [4]. Ivermectin has proven to be a safe and widely accepted treatment [8, 9], and is effective in reducing the filarial load [10, 11]. However, gaps remain in our knowledge regarding the effectiveness of a single round of triple-drug MDA in populations after an extended time-period. Evidence from field trials conducted in Papua New Guinea have shown that triple-drug MDA was more effective than two-drug MDA in reducing microfilariae (Mf) prevalence [8, 10], although a single round of triple-drug MDA was not sufficient to interrupt transmission [10]. Another study in Fiji observed no difference between triple-drug and two-drug MDA in a diurnally sub-periodic filarial transmission setting after 12 months [12].

The primary surveillance tool for determining whether MDA can be stopped is school-based Transmission Assessment Surveys (TAS) of children aged 6-7 years[1]. Thresholds for passing a TAS are parasite and vector specific. For *Wuchereria bancrofti* transmitted through an *Aedes* vector, passing a TAS requires the number of Ag-positive children identified in a survey to be below a given threshold, calculated so that the probability of an evaluation unit passing is at least 75% if the true Ag prevalence is 0.5%, and at most a 5% chance of falsely passing the TAS if the true Ag prevalence is ≥1% [1].

Antigenemia in younger children is more likely to be an indicator of recent infection, whilst antigenemia in older children and adults may be due to infections before MDA [1]. While sampling school children is considered convenient and cost-effective from a programmatic perspective, evidence suggests that Ag prevalence in younger children may not accurately reflect the Ag prevalence in the general population [13]. School-based TAS may therefore not be a suitable surveillance strategy for identifying ongoing LF transmission or determining elimination thresholds. Previous studies have shown that TAS can fail to detect hotspot areas of residual transmission, especially where prevalence is higher in older age groups, and may not be as sensitive an indicator of resurgence as older age groups [13, 14].

In Samoa, all evaluation units failed a school-based TAS in 2017. This led to the roll-out of a nationwide triple-drug MDA in 2018 from 14-26 August, the first such intervention to be implemented by any country at a national level [15]. The decision to use a triple-drug MDA was based on Samoa’s long history of endemic LF, with multiple rounds of two-drug MDA under the Pacific Programme for Elimination of LF (PacELF) between 1998 and 2017 proving insufficient for LF elimination [14]. The 2018 triple-drug MDA achieved good coverage, with an estimated 80% of the population taking the medication [15].

There were two main aims of this study. First, to assess impact of one round of triple-drug MDA against the previously reported baseline Ag prevalence [16]). Second, to assess the spatial epidemiology of LF in Samoa post triple-drug MDA. The specific objectives were to:

i. Report on change in Ag prevalence from 2018 to 2019, 6-8 months post triple-drug MDA.
ii. Investigate Mf prevalence and spatial epidemiology of LF in Samoa 6-8 months post triple-drug MDA
iii. Compare Ag prevalence between age groups (age 5-9 years vs age ≥10 years), sex (males vs females), and randomly vs purposively selected PSUs
iv. Compare geographic clustering of Ag positivity between 2018 and 2019, and between randomly and purposively selected PSUs.

This work expands on previously reported results [16, 17] by including a more extensive dataset from 35 PSUs compared to the 28 reported in McPherson et. al. (2022) [17], and detailed analysis of 2019 Ag prevalence by age, sex and PSU selection (random vs purposive), including analysis of the change in clustering of Ag positivity. The prevalence and epidemiology of the previously unreported 2019 Mf observations are also included.

## Methods

### Study location

Samoa is a Pacific Island nation with a population of around 200,000 [18]. It has a tropical climate with average rainfall of 3,000-6,000 mm/year [19]. There are two main islands, divided into four administration regions: three on the island of Upolu (Apia Urban Area [AUA], Northwest Upolu [NWU] and Rest of Upolu [ROU]) and the fourth being the island of Savai’i (SAV). The country is predominately rural with urban centres around Apia on Upolu and Salelolonga on Savai’i. The main vector associated with LF is *Aedes polynesiensis*, a day-biting mosquito that transmits the *W. bancrofti* parasite. In Samoa, the parasite is diurnally sub-periodic meaning that Mf circulate in the peripheral blood at any time but in higher concentrations during the daytime [20].

### Study design

The first baseline survey of 35 primary sampling units (PSUs) was conducted from 26 September to 9 November 2018, with all participants enrolled within two months from the start of the first round of triple-drug MDA. Thirty PSUs were randomly selected and the remaining five PSUs were purposively selected by the Samoa Ministry of Health as suspected hotspots based on results of previous surveys. Participant recruitment and target sample sizes have been described in detail elsewhere [16]. Briefly, from each PSU, we aimed to recruit 57 participants ages ≥10 years and 57 aged participants 5-9 years. Sample sizes were calculated to detect 2% Ag prevalence in each age group, with a 5% chance of type 1 error, 75% power (when true prevalence is 1%), and a design effect of 2.0.

Fifteen households were selected per PSU using a virtual walk method [16]. Each household was visited at least once between 3pm and 8pm, Monday to Saturday. If the selected building was not a house, or the residents declined to participate, it was replaced with the nearest household. If no one was at home, field teams made a second visit where possible. If the target sample size for participants aged ≥10 years was not reached after visiting every selected household at least once, houses where nobody was at home were replaced. If there were still insufficient participants after 15 households were surveyed, up to five additional households were randomly selected.

In each PSU, a convenience survey of 5-9-year-old children was conducted. Surveys took place within the PSU, generally at a community location such as a church, school, or household of a community leader. The convenience surveys were arranged in collaboration with the village mayor, and with the permission of the village Women’s Committee. If insufficient children were recruited after the first visit, either a second session was organised, or the field teams randomly selected additional households within the village to recruit participants in this age group.

### Data collection

Surveys were conducted by community workers from the Samoa Red Cross Society, working with the research team. Field teams collected the GPS coordinates of each household enrolled in the community surveys using smartphones. Demographic information was collected from each consenting participant using a standardised electronic questionnaire with the Standard Data kit (SDK) software (https://www.datastandard.co). A clinical examination was conducted for scabies (including residents of all ages), the results of which have been reported elsewhere [15]. During the convenience surveys, field teams collected demographic data from a child’s parent or guardian using a short electronic questionnaire. For all participants aged ≥5 years, a finger prick blood sample (up to 400 μL) was collected into a heparin microtainer.

### Sample processing

Blood samples were kept cool using ice packs until they were refrigerated upon field teams’ return to the field laboratory. Samples were processed within 48 hours of collection. Blood samples were brought back to room temperature before being tested for Ag using Alere® Filariasis Test Strips (FTS) (Scarborough, ME, USA), which were read at 10 minutes. Any blood samples that tested Ag-positive were used to prepare up to three thick blood slides, each with three 20μL lines of blood as per WHO guidelines [1]. Slides were dehaemoglobinised in water for 10-15 minutes and left to dry for 72 hours. Two of the three slides were fixed with 100% methanol and stained with Giemsa according to WHO-recommended methods [1] and read by independent readers (one slide per reader). A sample was considered Mf-positive if either reader identified Mf on any of the slides.

### Data analysis

Ag and Mf prevalence for 2018 and 2019 were estimated at national, regional and PSU levels. Estimates were calculated using the Stata (Stata Corp, TX, USA) *proportion* command for each age category (5-9 and ≥10 years) and adjusted for selection probability at PSU and household levels and clustering at PSU level. Results were standardised by age group and gender against matching 2016 census distributions and adjusted for survey design. Presence/absence of Ag and Mf were mapped at the PSU level. Mf/mL for each Mf-positive participant was calculated as the average number of Mf per 60 μL of blood observed on each slide, converted to Mf/mL. Mf density was calculated as the geometric mean (due to skewed distributions) of the Mf/mL density for all Mf-positive participants in 2019.

To examine changes in Ag prevalence over time at the national level, we fitted multivariable mixed effect logit models with random/group effects for each PSU at baseline (2018), and random/group effects for the temporal change at the PSU level. To examine changes over time by region, we modified the model to include fixed/population effects for each region in 2018 and fixed/population effects for the temporal change. From these models we calculated ORs and 95% confidence intervals (CIs) to determine significant changes in prevalence between years. Geographic clustering of Ag at the household, PSU, and regional levels were assessed using intra-cluster correlation (ICC), estimated using the same multi-level mixed effects logistic regression model formulation adjusting for age and sex, and compared between years. As the 2018 survey was conducted post-MDA a comparison of Mf prevalence between 2018 and 2019 was not appropriate.

### Ethics

Ethics approval was obtained from Human Research Ethics Committees at the Samoa Ministry of Health and The Australian National University (protocol 2018/341) and ratified by the University of Queensland Human Research Ethics Committee (protocol 2021/HE000895). This work was conducted in close collaboration with the Samoa Ministry of Health, the WHO country office in Samoa, and the Samoa Red Cross. Culturally appropriate methods for survey design and implementation were considered at each stage of the study. Before entering a village, consent was obtained from village leaders for their community’s involvement. Village leaders were also asked to inform community members about the study prior to the field team’s visits. On arrival at each selected household, a senior field team member approached the household to give a verbal and written explanation of the study to an adult resident and sought verbal approval to enter the household.

Written informed consent (and verbal assent from minors) was obtained from each individual (or a parent/guardian of child participants) prior to enrolment in the study. Written information, consent forms, and surveys were provided/conducted in Samoan or English according to each participant’s preference. Local bilingual field teams carried out all field activities within the villages in a culturally sensitive manner.

## Results

### Demographics of study population

The demographic characteristics of the study population in 2018 [16] and 2019 are summarised in Table **1**. In 2019, 4290 participants were recruited across the household (n=2629) and convenience surveys (n=1661). Mean age was 20.3 years (range: 5-89) and 52% of participants were female. A total of 3654 participants were recruited from the 30 randomly selected PSUs, and 636 from the five purposively selected PSUs. The mean number of households per PSU was similar between randomly selected (15.7) and purposively selected PSUs (15.0).

**Table 1.**
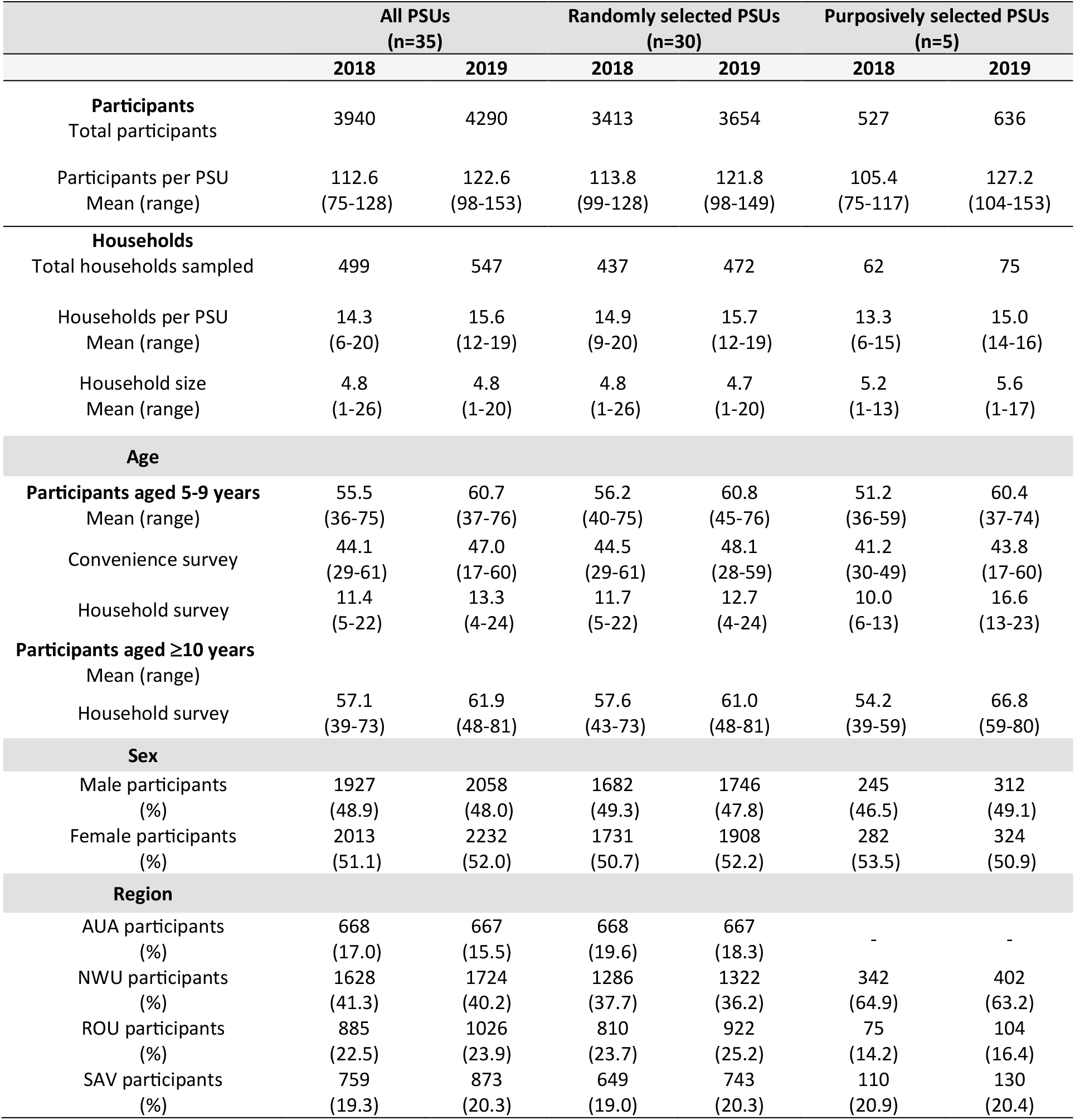
Sample size and demographic characteristics of study population in the 2018 survey (1.5 to 3.5 months post triple-drug MDA) [16] and 2019 survey (six to eight-months post triple-drug MDA). Results are presented for all 35 primary sampling units (PSUs), and separately for randomly and purposively selected PSUs as well as combined.

### Adjusted antigen prevalence in 2019

Of the 4290 participants in 2019, a valid FTS result was available for 4256 (99.2%) participants, of which 139 (3.3%) were Ag-positive. Ag-positive PSUs (PSUs with at least one Ag-positive participant) were identified in all four regions (Fig 1). Adjusted Ag prevalence in the 30 randomly selected PSUs was 4.1% (95% CIs 2.7-5.9%) and there was significantly higher Ag-prevalence in purposively selected PSUs (14.9%, 95% CIs 13.7-16.0%; *P*<0.001). Patterns of Ag-positivity in the random PSUs were similar to those observed in 2018; significantly higher Ag prevalence was seen in participants aged ≥10 years vs 5-9 years (4.6%, 95% CIs 3.0-6.7% vs 1.1%, 95% CIs 0.5-2.2%; *P*<0.001), in males vs females (6.7%, 95% CIs 4.4-9.8% vs 1.3%, 95% CIs 0.5-2.7%; *P*<0.001). For the 30 PSUs, Ag prevalence was highest in SAV (10.2%) and lowest in AUA (1.5%). Full details of Ag prevalence, including by region, are provided in Supplementary S1 Table and S2 Table.

**Fig 1.**
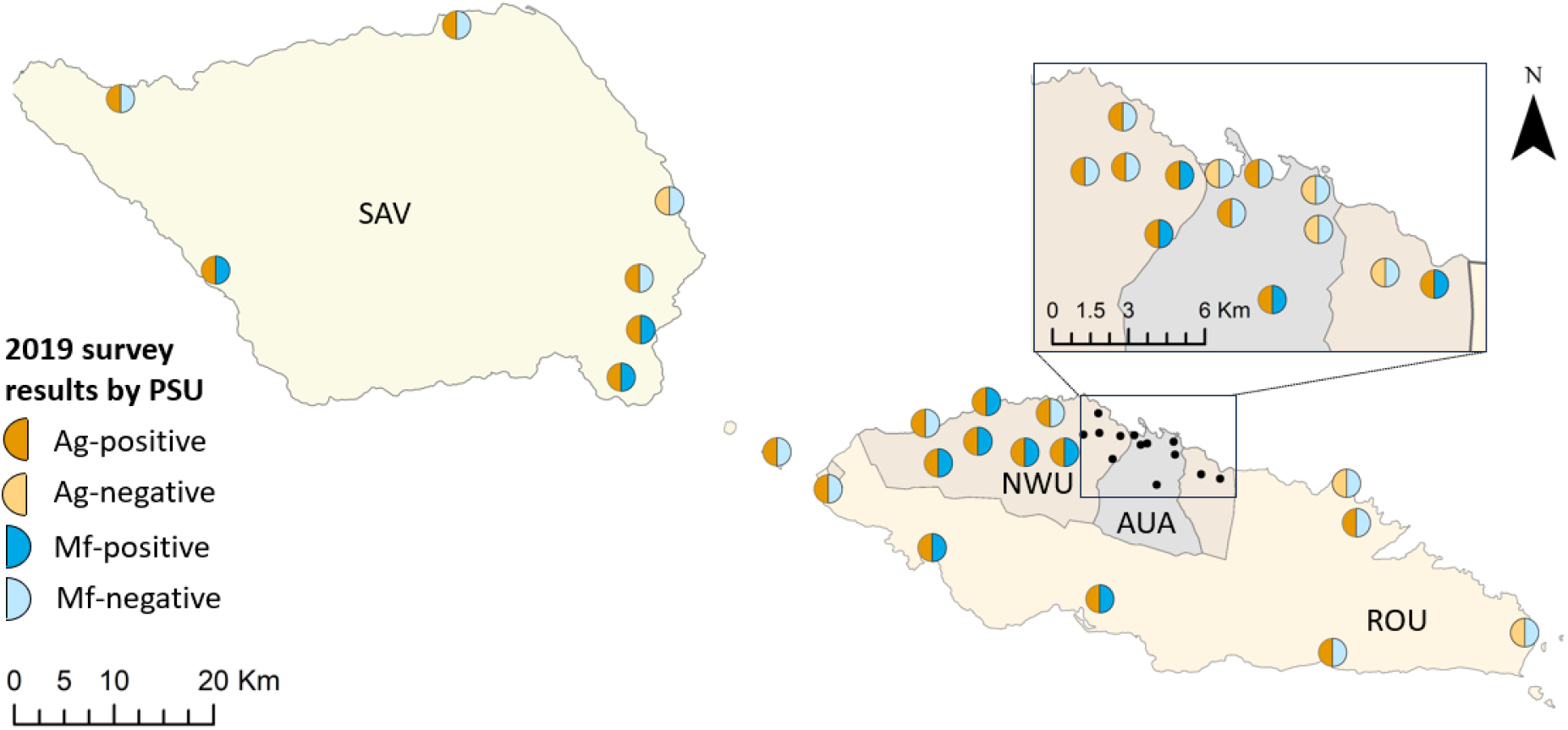
Observed Ag and Mf infection at the PSU level in Samoa in 2019. Regions shown are Apia urban area (AUA), Northwest Upolu (NWU), Rest of Upolu (ROU) and Savai’i (SAV). Spatial data on country, island, region, and village boundaries in Samoa were obtained from the Pacific Data Hub (pacificdata.org accessed on 8 July 2020) and DIVA-GIS (diva-gis.org, accessed on 12 August 2019) under an open access licence available at https://pacific-data.sprep.org/resource/public-data-license-agreement-0.

### Adjusted microfilaria prevalence in 2019

In 2019, 32 (0.8%) participants tested Mf-positive. Mf-positive PSUs (PSUs with at least one Mf-positive participant) were identified in all four regions (Fig 1). Adjusted Mf prevalence in the 30 randomly selected PSUs was 0.8% (95% CIs 0.3-1.6%), with an average Mf density (geometric mean) among Mf-positive participants of 138.7 Mf/mL (min= 8.3, max= 2558.3, median= 170.8, IQR=250.0). A significantly higher prevalence of Mf-positives was found in purposively selected vs randomly selected PSUs (4.3%, 95% CIs 3.4-5.4% vs 0.8%, 95% CIs 0.3-1.6% ; *P*<0.001), in participants aged ≥10 years vs those aged 5-9 years (0.9%, 95% CIs 0.3-1.8% vs 0.1% 95% CIs 0.0-0.8; *P*=0.045), and in male vs females (1.3%, 95% CIs 0.5-2.8% vs 0.3%, 95% CIs 0.1-0.8; *P*=0.007). Full details of Mf prevalence, including by region, are provided in Supplementary S2 Table and S3 Table.

### Change in Ag from 2018 to 2019

In the randomly selected PSUs, there was no change in the adjusted Ag prevalence from 2018 (3.9%) to 2019 (4.1%). By age group, Ag prevalence was unchanged in 5-9-year-olds from 2018 to 2019 (1.2% vs 1.1%; *P*=0.793) and in ≥10-year-olds (4.7% vs 4.1%; *P*=0.547) (Figure 2, panel C). There was non-significant increase in Ag prevalence among male participants between 2018 and 2019 (4.7vs 6.7%; *P*=0.361), and a non-significant decrease among female participants (3.1% vs 1.3%; *P*=0.052). Confidence intervals are reported in Supplementary S1 Table.

**Fig 2.**
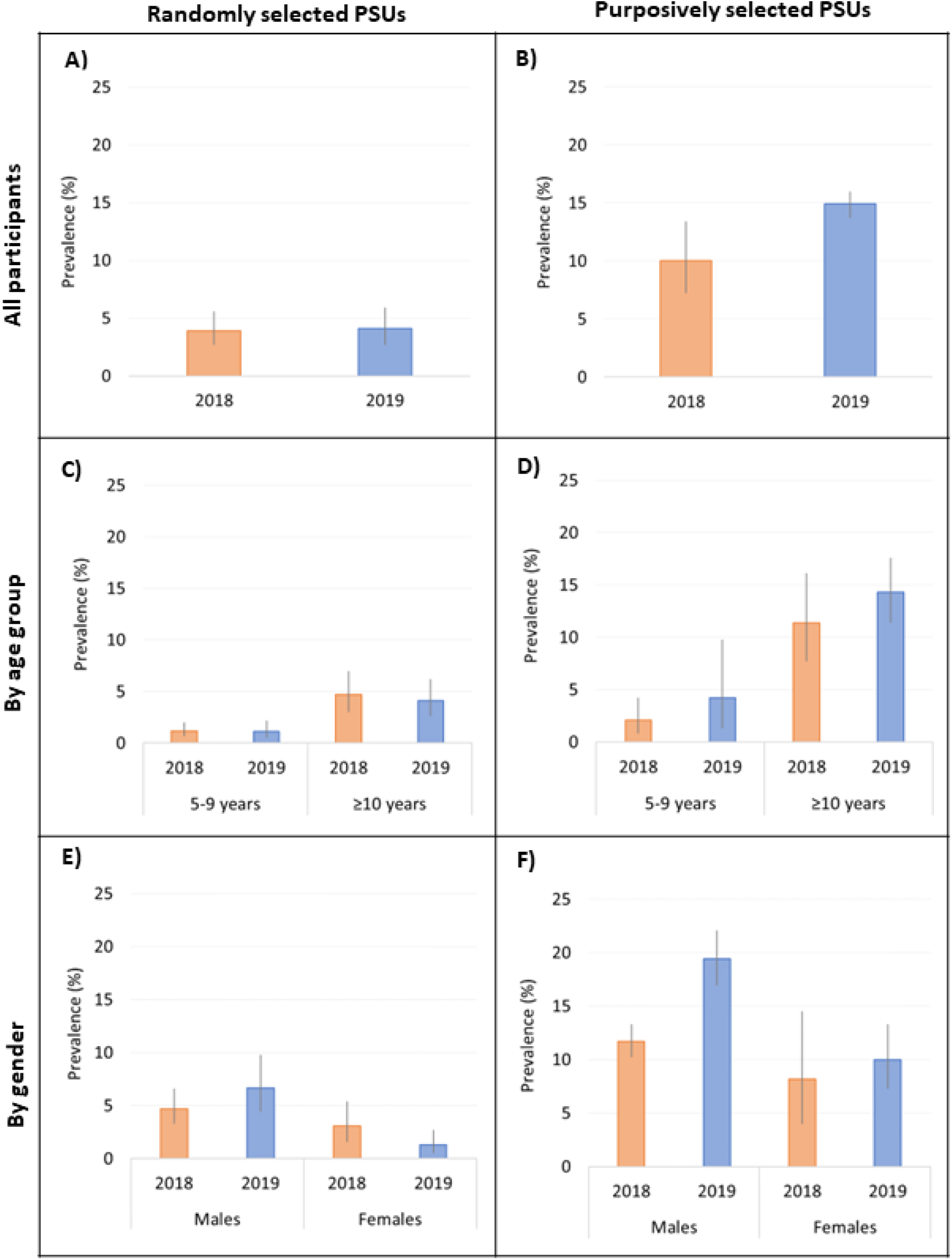
Adjusted antigen prevalence in randomly selected and purposively selected PSUs in 2018 [orange] and 2019 [blue] for all participants (A,B), and stratified by age (C, D) and sex (E,F).

**Fig 3.**
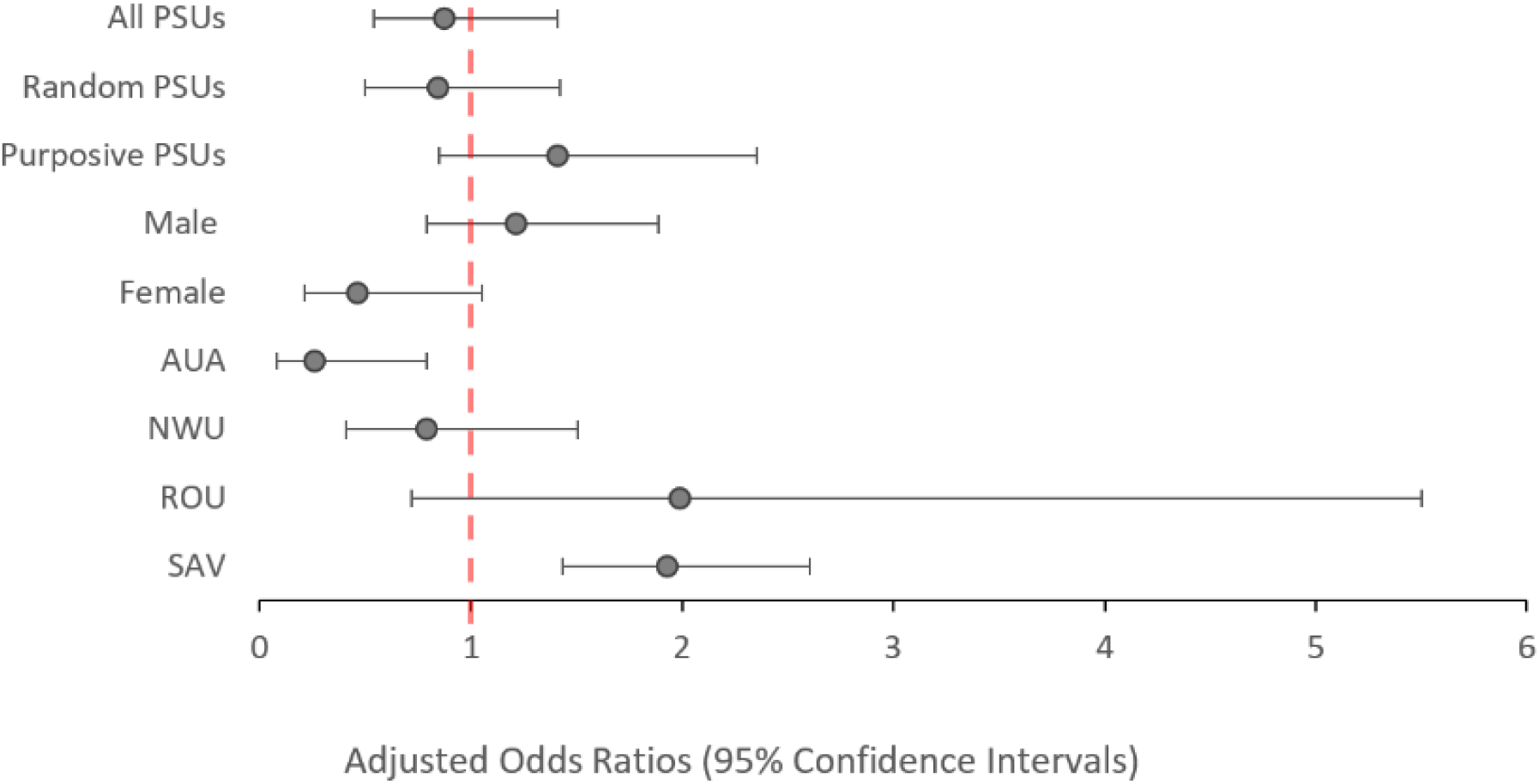
Adjusted odds ratios of testing positive to antigen in 2019 compared to 2018 (reference value) among all participants aged ≥5 years stratified by PSU selection, sex and region. Values above 1.0 indicate and increased odds of testing antigen-positive. Values below 1.0 indicate a reduced odds of testing antigen-positive.

In purposively selected PSUs, the adjusted Ag prevalence did not change significantly from 2018 (10.0%) to 2019 (14.9%; Fig 2, panels A and B). By age group, Ag prevalence was not significantly different. among 5-9-year-olds (2.1% vs 4.2%; *P*=0.074) and ≥10-year-olds (11.4% vs 14.3%; *P*=0.200) between 2018 and 2019 (Fig 2, panels C and D). Between 2018 and 2019, Ag prevalence for male participants (11.7% vs 19.4%; *P*=0.077) and female participants (8.2% vs 10.0%; *P*=0.234) did not change significantly (Fig 2, panels E and F). Confidence intervals (95%) are reported in Supplementary S3 Table.

### Odds of Ag positivity by year

There was no significant change in the odds of testing Ag-positive in 2019 compared to 2018 (aOR: 0.87; *P*=0.573), irrespective of whether participants were from purposively selected PSUs (aOR: 1.41; *P*=0.135) or randomly selected PSUs (aOR: 0.84; *P*=0.498). At the regional level, participants from AUA had significantly lower odds of testing Ag-positive in 2019 vs 2018 (aOR: 0.26; *P*=0.026), while participants from SAV had significantly increased odds of testing Ag-positive in 2019 vs 2018 (aOR: 1.93; *P*=0.002). Confidence intervals (95%) are reported in Supplementary S2 Table and S3 Table.

When all 35 PSUs were considered, there was no significant change in the odds of testing Ag-positive between 2018 and 2019 among participants aged 5-9 years (aOR=0.95; *P*=0.894) (Fig 4) or among participants aged ≥10 years (aOR: 0.86; *P*=0.573). However, participants aged ≥10-years from AUA had significantly lower odds (aOR: 0.28; *P*=0.033), while those from SAV had significantly higher odds (aOR: 1.96; *P*=0.003) of testing Ag-positive in 2019 vs 2018 (Fig 5).

**Fig 4.**
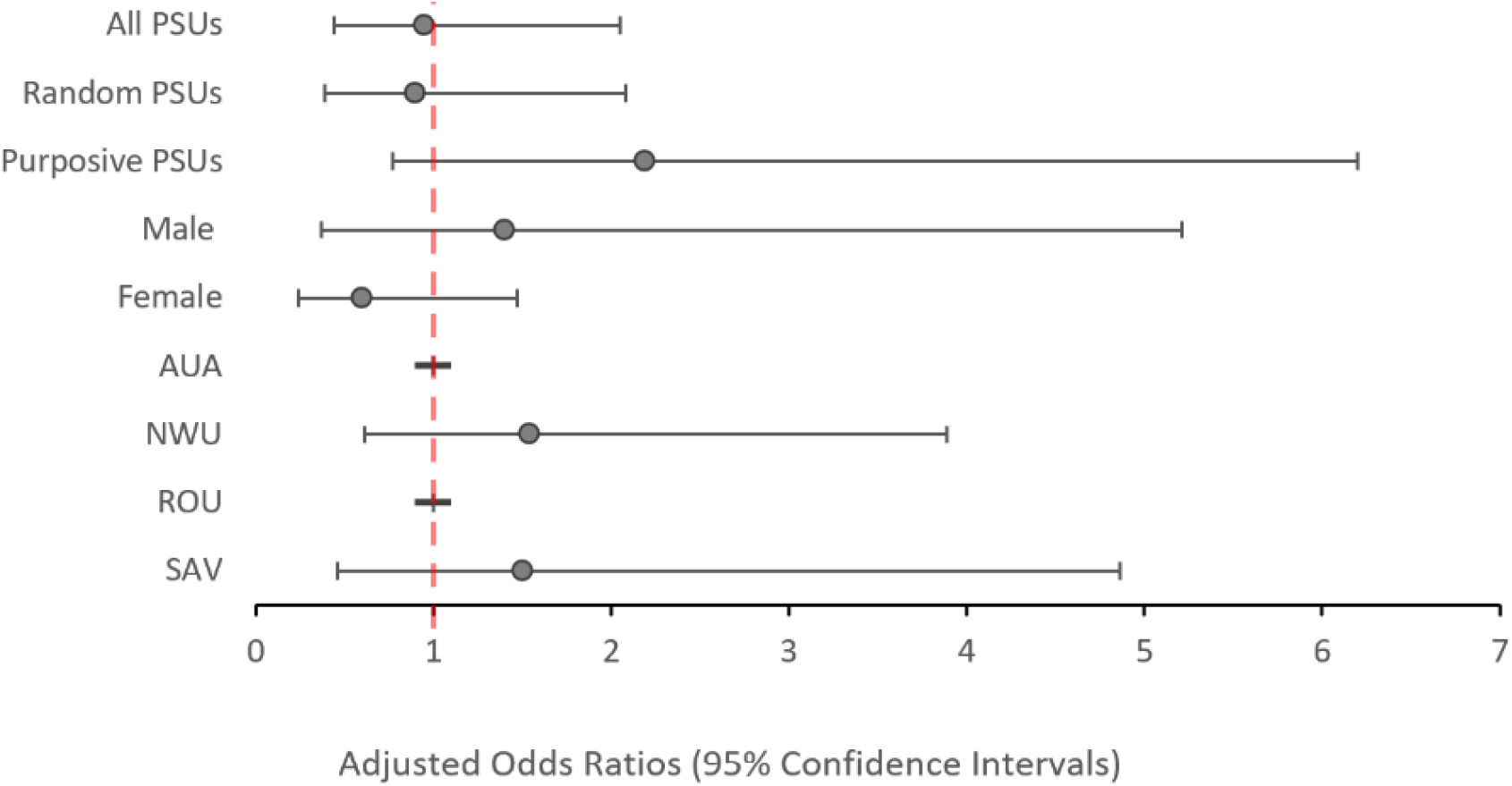
Adjusted odds ratios of testing positive to antigen in 2019 compared to 2018 (reference value) among all participants aged 5-9 years stratified by PSU selection, sex and region. Values above 1.0 indicate an increased odds of testing antigen-positive. Values below 1.0 indicate a reduced odds of testing antigen-positive. There were no Ag-positive cases in AUA or ROU in 2019 for this age group.

**Fig 5.**
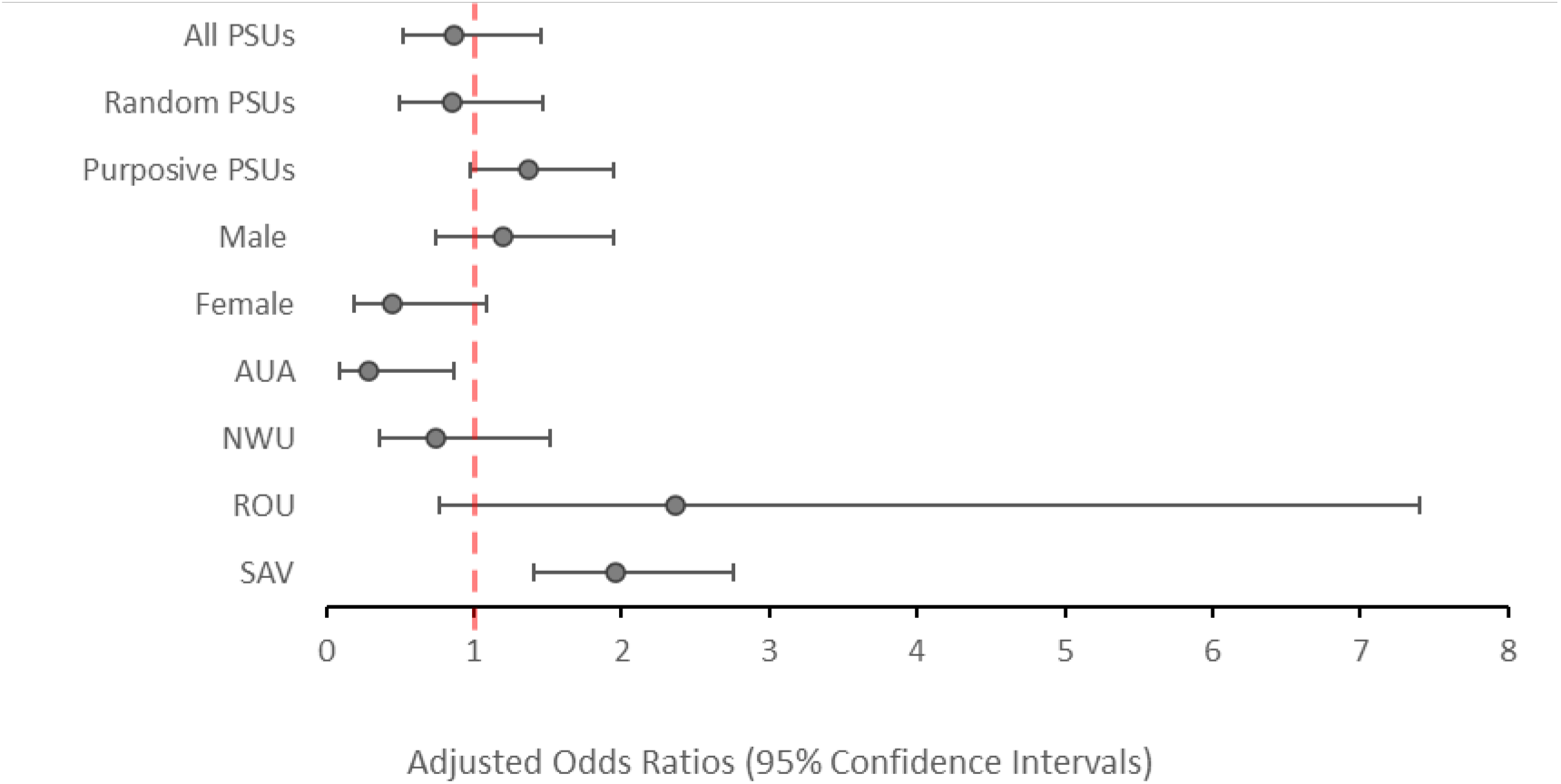
Adjusted odds ratios of testing positive to antigen in 2019 compared to 2018 (reference value) among all participants aged ≥10 years stratified by PSU selection, sex and region. Values above 1.0 indicate and increased odds of testing antigen-positive. Values below 1.0 indicate a reduced odds of testing antigen-positive.

### Clustering of Ag participants

As with the 2018 survey, ICC of Ag-positive participants across the 35 PSUs in 2019 was highest at the household level (ICC 0.51), suggesting a high degree of clustering within households compared to the PSU (ICC 0.23), or regional level (ICC 0.02). There was no significant change in clustering between years at PSU, regional or household levels. These results were consistent for both the 30 randomly and five purposively selected PSUs, as well as when calculated for the combined 35 PSUs (Supplementary S4 Table).

## Discussion

Ag prevalence in Samoa had not fallen to below 2% within 6-8 months after the first round of triple-drug MDA, which was distributed in 2018. Results show evidence of ongoing transmission, indicating that further intervention is needed to achieve elimination. This is in line with the current WHO recommendations that advocate multiple rounds of MDA [4]. Ag prevalence was lower in the younger vs older age group (aged 5-9 vs ≥10 years) and in the randomly selected vs purposively selected PSUs.

Of the four regions, only AUA had statistically significant lower odds of testing Ag-positive in 2019 vs 2018, despite having the lowest reported coverage in the 2018 MDA [15]. AUA is highly urbanised compared to the other three regions, which could contribute to lower levels of transmission due to environmental factors, but this link remains to be explored. The low baseline for Ag prevalence in the AUA PSUs in 2018 also means that a larger relative difference is achieved for a similar absolute reduction in infection, i.e., when very few cases are observed a small number of additional cases will have a large effect on the percent increase/decrease in prevalence.

In 2019, Ag prevalence remained lower in participants aged 5-9-years compared to participants aged ≥10 years. This is consistent with existing evidence of higher Ag prevalence in older age groups in the Pacific region [16, 21]. Higher Ag and Mf prevalence in adults supports the notion that prevalence measured in school-based TAS may not be an appropriate estimate to extrapolate to the general population, as 6-7 year old children are less likely to be representative of the overall population.

Targeted sampling to efficiently find and treat foci of residual infection has been suggested as a complementary intervention to widescale MDA [22, 23]. Targeted strategies that rely on reactive case finding (also referred to as snowball sampling) assume there is a high chance that infected people will be found near other infected people, i.e. that infections are clustered. The results presented here support previous findings in this area [24]and suggest that targeted sampling of household members, and potentially near neighbours would be an efficient strategy for locating LF infections in people.

Further investigation is needed to better understand the potential efficiency gains from targeted sampling for on-the-ground LF surveillance. Similarly, the findings from this study that MDA may have been less effective in reducing Ag prevalence in males than in females, particularly in the older age group, suggests that post-MDA follow-up surveillance should target this demographic for locating and treating residual infection.

Results from a concurrent survey using molecular xenomonitoring (MX) of mosquitoes demonstrated that in Samoa, the 2018 MDA decreased LF infection prevalence in mosquitoes in the short term [17]. Given the short time between the human surveys in 2018 and 2019 (approximately five to six months), it is possible that a there was a reduction in active LF infections in humans, but insufficient time had passed to allow the corresponding drop in Ag to be detected. The timing of the surveys will also have underestimated any change in Mf from 2018 to 2019, as the first survey took place shortly after the triple-drug MDA. While this would not be expected to have any impact on Ag levels, Mf are cleared quickly following treatment [25] and the 2018 Mf prevalence presented is not an accurate baseline (Supplementary S1 Table). A comparison of Mf prevalence between the two years has therefore not been included here.

Our results show that a single round of triple-drug MDA was insufficient to break transmission of the LF parasite. Although evidence published elsewhere attests to the efficacy of a single dose of the triple-drug regime at clearing Mf at the individual level [8, 25], further MDA rounds are needed to reduce infection levels sufficiently to break transmission at a population level. Higher Ag and Mf prevalence in those aged ≥10 years, and the presence of substantial clustering of Ag at the household level suggests that targeted surveillance strategies for these groups are needed to support LF elimination pro**grams**.

## Data Availability

Data used in this paper were collected during field surveys in Samoa. Communities in Samoa are small (some with less than 200 inhabitants) and sharing individual level data could enable identification of individual participants, and violating the conditions of the study’s ethics approval. For requests relating to data access, please contact the Human Ethics Department at the University of Queensland citing protocol 2021/HE000895. All relevant data at the primary sampling unit level has been included in the supplementary material.

## Acknowledgements

We would like to thank all the staff at the Samoa Ministry of Health who supported the many different aspects of the study. We especially thank Miriama Asoiva who provided valuable advice on local logistics and cultural sensitivities, and assistance with obtaining permissions to conduct village visits. We also thank Tile Ah Leong-Lui, Fuatai Maiava and Siatua Loau for sharing their knowledge about the LF elimination program in Samoa, and to Fuatai for making it possible for nurses to assist with the household surveys. We sincerely thank Tautala Maula, the general secretary of the Samoa Red Cross, and her team (especially Babey Suniula, Nixon Mataia, Brenda Koon Wai You, Alesi Mataia, and Shem Lepale) for their enthusiastic and untiring support with fieldwork, village visits, and laboratory work; this survey would not have been possible without their hard work and dedication. We greatly appreciate all the support and advice provided by Rasul Baghirov and Lepaitai Hansell at the WHO country office in Samoa, and thank them for generously sharing their wisdom. We also thank the Australian volunteers and students (Gabriela Willis, Meru Sheel, Benjamin Dickson, Brady McPherson, Kelley Meder and Kei Owada) who assisted with fieldwork and data management, and technical advice provided by Patrick Lammie (Task Force for Global Health) and Kimberly Won (US Centers for Disease Control and Prevention). We thank the NIH/NIAID Filariasis Research Reagent Resource Center (www.filariasiscenter.org) for supplying positive controls for the Filariasis Test Strips

## Supporting Information Captions

**Supplementary S1 Table**. Ag and Mf prevalence in 30 randomly selected primary samples units (PSUs) in Samoa in 2018 (1.5-3.5 months post triple-drug MDA) and 2019 (6-8 months post triple-drug MDA).

**Supplementary S2 Table**. Ag and Mf prevalence in 35 primary sampling units (PSUs) in 2019 in Samoa. Standardised by age and gender and adjusted for survey design.

**Supplementary S3 Table**. Ag and Mf prevalence in 5 purposively selected primary samples units (PSUs) in Samoa in 2018 (1.5-3.5 months post triple-drug MDA) and 2019 (6-8 months post triple-drug MDA).

**Supplementary S4 Table**. Clustering of Ag-positive participants in Samoa in 2018 and 2019 at the regional, PSU and household level

## References

1. World Health Organization. Monitoring and Epidemiological Assessment of Mass Drug Administration in the Global Programme to Eliminate Lymphatic Filariasis: A Manual for National Elimination Programmes. Geneva, Switzerland: 2011.

2. World Health Organization. Ending the neglect to attain the Sustainable Development Goals: a road map for neglected tropical diseases 2021–2030. 2020.

3. Turner HC, Ottesen EA, Bradley MH. A refined and updated health impact assessment of the Global Programme to Eliminate Lymphatic Filariasis (2000–2020). Parasites & Vectors. 2022;15(1):181. doi: 10.1186/s13071-022-05268-w.

4. World Health Organization. Guideline: Alternative Mass Drug Administrations to Eliminate Lymphatic Filariasis Geneva, Switzerland:2017 [19/09/2023]. Available from: http://apps.who.int/iris/bitstream/handle/10665/259381/9789241550161-eng.pdf.

5. World Health Organization. WHO officially recognizes noma as a neglected tropical disease Geneva2023. Available from: https://www.who.int/news/item/15-12-2023-who-officially-recognizes-noma-as-a-neglected-tropical-disease.

6. The roadmap towards elimination of lymphatic filariasis by 2030: insights from quantitative and mathematical modelling. Gates Open Res. 2019;3:1538. Epub 2019/11/16. doi: 10.12688/gatesopenres.13065.1. PubMed PMID: 31728440; PubMed Central PMCID: PMCPMC6833911.

7. World Health Organisation. Global Health Observatory - Lymphatic filariasis (Elephantiasis) 2023 [cited 2024]. Available from: https://www.who.int/data/gho/data/themes/topics/lymphatic-filariasis.

8. Tavul L, Laman M, Howard C, Kotty B, Samuel A, Bjerum C, et al. Safety and efficacy of mass drug administration with a single-dose triple-drug regimen of albendazole + diethylcarbamazine + ivermectin for lymphatic filariasis in Papua New Guinea: An open-label, cluster-randomised trial. PLoS Negl Trop Dis. 2022;16(2):e0010096. Epub 20220209. doi: 10.1371/journal.pntd.0010096. PubMed PMID: 35139070; PubMed Central PMCID: PMCPMC8863226.

9. Weil GJ, Bogus J, Christian M, Dubray C, Djuardi Y, Fischer PU, et al. The safety of double- and triple-drug community mass drug administration for lymphatic filariasis: A multicenter, open-label, cluster-randomized study. PLoS Med. 2019;16(6):e1002839. Epub 20190624. doi: 10.1371/journal.pmed.1002839. PubMed PMID: 31233507; PubMed Central PMCID: PMCPMC6590784.

10. Laman M, Tavul L, Karl S, Kotty B, Kerry Z, Kumai S, et al. Mass drug administration of ivermectin, diethylcarbamazine, plus albendazole compared with diethylcarbamazine plus albendazole for reduction of lymphatic filariasis endemicity in Papua New Guinea: a cluster-randomised trial. Lancet Infect Dis. 2022;22(8):1200–9. Epub 20220506. doi: 10.1016/s1473-3099(22)00026-3. PubMed PMID: 35533701; PubMed Central PMCID: PMCPMC9300473.

11. Biritwum N-K, Yikpotey P, Marfo BK, Odoom S, Mensah EO, Asiedu O, et al. Persistent ‘hotspots’ of lymphatic filariasis microfilaraemia despite 14 years of mass drug administration in Ghana. Transactions of The Royal Society of Tropical Medicine and Hygiene. 2017;110(12):690–5. doi: 10.1093/trstmh/trx007.

12. Hardy M, Samuela J, Kama M, Tuicakau M, Romani L, Whitfeld MJ, et al. Individual Efficacy and Community Impact of Ivermectin, Diethylcarbamazine, and Albendazole Mass Drug Administration for Lymphatic Filariasis Control in Fiji: A Cluster Randomized Trial. Clin Infect Dis. 2021;73(6):994–1002. doi: 10.1093/cid/ciab202. PubMed PMID: 33728462.

13. Sheel M, Sheridan S, Gass K, Won K, Fuimaono S, Kirk M, et al. Identifying residual transmission of lymphatic filariasis after mass drug administration: Comparing school-based versus community-based surveillance - American Samoa, 2016. PLOS Neglected Tropical Diseases. 2018;12(7):e0006583. doi: 10.1371/journal.pntd.0006583.

14. Graves PM, Joseph H, Coutts SP, Mayfield HJ, Maiava F, Ah Leong-Lui TA, et al. Control and elimination of lymphatic filariasis in Oceania: Prevalence, geographical distribution, mass drug administration, and surveillance in Samoa, 1998-2017. Advances in parasitology. 2021;114:27–73. Epub 2021/10/27. doi: 10.1016/bs.apar.2021.03.002. PubMed PMID: 34696844.

15. Willis GA, Mayfield HJ, Kearns T, Naseri T, Thomsen R, Gass K, et al. A community survey of coverage and adverse events following country-wide triple-drug mass drug administration for lymphatic filariasis elimination, Samoa 2018. PLOS Neglected Tropical Diseases. 2020;14(11):e0008854. doi: 10.1371/journal.pntd.0008854.

16. Lau CL, Meder K, Mayfield HJ, Kearns T, McPherson B, Naseri T, et al. Lymphatic filariasis epidemiology in Samoa in 2018: Geographic clustering and higher antigen prevalence in older age groups. PLOS Neglected Tropical Diseases. 2020;14(12):e0008927. doi: 10.1371/journal.pntd.0008927.

17. McPherson B, Mayfield HJ, McLure A, Gass K, Naseri T, Thomsen R, et al. Evaluating Molecular Xenomonitoring as a Tool for Lymphatic Filariasis Surveillance in Samoa, 2018–2019. Tropical Medicine and Infectious Disease. 2022;7(8):203. PubMed PMID: doi:10.3390/tropicalmed7080203.

18. Samoa Bureau of Statistics. Samoa population and housing census 2021 basic tables. Apia, Samoa: 2022.

19. Government of Samoa Meteorology Division. Climatology 2020 [cited 2024]. Available from: http://www.samet.gov.ws/index.php/climatology.

20. Ramalingam S. The epidemiology of filarial transmission in Samoa and Tonga. Ann Trop Med Parasitol. 1968;62(3):305–24. doi: 10.1080/00034983.1968.11686565. PubMed PMID: 5751950.

21. Lau CL, Sheel M, Gass K, Fuimaono S, David MC, Won KY, et al. Potential strategies for strengthening surveillance of lymphatic filariasis in American Samoa after mass drug administration: Reducing ‘number needed to test’ by targeting older age groups, hotspots, and household members of infected persons. PLOS Neglected Tropical Diseases. 2021;14(12):e0008916. doi: 10.1371/journal.pntd.0008916.

22. Mayfield HJ, Sturrock H, Arnold BF, Andrade-Pacheco R, Kearns T, Graves P, et al. Supporting elimination of lymphatic filariasis in Samoa by predicting locations of residual infection using machine learning and geostatistics. Scientific Reports. 2020;10(1):20570. doi: 10.1038/s41598-020-77519-8.

23. Riches N, Badia-Rius X, Mzilahowa T, Kelly-Hope LA. A systematic review of alternative surveillance approaches for lymphatic filariasis in low prevalence settings: Implications for post-validation settings. PLOS Neglected Tropical Diseases. 2020;14(5):e0008289. doi: 10.1371/journal.pntd.0008289.

24. Shaw C, McLure A, Graves PM, Lau CL, Glass K. Lymphatic filariasis endgame strategies: Using GEOFIL to model mass drug administration and targeted surveillance and treatment strategies in American Samoa. PLOS Neglected Tropical Diseases. 2023;17(5):e0011347. doi: 10.1371/journal.pntd.0011347.

25. Graves PM, Sheridan S, Scott J, Amosa-Lei Sam F, Naseri T, Thomsen R, et al. Triple-Drug Treatment Is Effective for Lymphatic Filariasis Microfilaria Clearance in Samoa. Tropical Medicine and Infectious Disease. 2021;6(2):44. PubMed PMID: doi:10.3390/tropicalmed6020044.

